# OPTIMIZING COVID-19 VACCINE USAGE

**DOI:** 10.1101/2021.01.04.21249167

**Authors:** Carlos Poblete Jara, Lício A. Velloso, Eliana Pereira de Araújo

## Abstract

As the worldwide vaccination, it is imperative to minimize vaccine wastage by effectively using all doses available. Vaccine wastage can occur at multiple points during the vaccination process, but it is mainly because the device dead space and the filling process technique. However, there are no studies discussing the waste volume effect of COVID-19 vaccines in clinical practice. There is an increasing COVID-19 vaccine demand that we estimate up to several billion dual doses. The objective of this study was to assess the number of 0.3mL doses obtained from a multiple-dose vial using 1ml and 3ml syringes with different type of needles replicating the first COVID-19 vaccination protocol.

Our results suggest that it is possible to obtain six or seven doses from each vial instead five. We provide evidence to optimize between 20% and 40% additional vaccine doses per vial if the current 5-dose vials are used, making scarce supplies go further.

It is our duty, as researchers, to ensure the efficacy and efficiency of the worldwide COVID-19 vaccination process. However, if standard syringes-needles and technique are used, there may not be sufficient volume to draw extra doses from a single vial.

## INTRODUCTION

SARS-CoV-2, the pathogenic agent of COVID-19, has had devastating consequences globally. Worldwide, COVID-19 overwhelm lifestyle, macroeconomic and threatens to affect billion human lives ^1-3^.

The COVID-19 vaccination campaigns are being conducted to interrupt SARS-CoV-2 transmission and has shown efficacy in preventing COVID-19 illness^4-9^. Vaccines are needed to prevent COVID-19 and to protect people who are at high risk for complications ^4^. Currently, there are only few companies producing COVID-19 vaccines, and optimization of usage could expand considerably the number of people immunized. There are currently not many manufacturers globally of COVID-19 vaccines to provide a short period proper number of vaccines for the world population. In this way, it is imperative to avoid vaccine wastages by effectively use all doses available. Vaccine wastage can occur at multiple points during the preparation, filling, and inoculation processes, but it is mainly because the device dead space and filling process technique^10-15^:

One vital device is an appropriate syringe-needle system for delivering the vaccine. The association between needle-syringe system and waste were previously reported^10,16,17^. However, there are no publications discussing the waste volume effect of COVID-19 vaccines in clinical practice.

With the possibility of billions to be infected and the limited vaccine supply, it is essential that the syringe-needle system used waste as little as possible and consequently allow for a higher number of people vaccinated^17^.

Here, we assessed the number of doses draw from a multiple-dose vial, simulating the first COVID-19 vaccination protocol. Our goal was to optimize the COVID-19 vaccine wastage. We showed different syringe-needle system to use and we explored a no dead-volume approach to draw a seventh dose.

## RESULTS

### Syringe – needle system wastage

We simulated the vaccination process pre-loading 0.45mL of saline diluted with 1.8 mL of 0.9% Sodium Chloride (Figure 1a). We found the mean dead-volume of the syringe – needle system ranged from 40 μL to 560 μL per vial (Table 1). The lower dead-volume was found in the no dead-volume group (Figure 1b, e) and the higher waste volume in the 3 ml syringe-23Gx1 needle group (Figure 1c, e). Also, we found the aspiration varied between needle Gauge and syringe system and ranged from 1.69 mL to 2.21 mL per vial (Table 1). Specifically, our results showed that the 3 mL syringe attached to 27Gx1/2 needle, increased the total volume recovered from the multiple-dose vial compared to the 3 mL syringe-23Gx1 needle system (Table 1, Figure d).

**Table 1.** Recovered: mean value of fluid withdraws from each vial. Dead volume: mean value of the difference between the 2.25 mL initial load and the recovered fluid from each vial. Doses: mean value of number of doses from each vial. % > 5 doses, % > 6 doses, and % > 7 doses: percentage of times reaching the pointed value. (a): no dead-volume approach. Mean values± Standard deviation. All experiments in quadruplicate.

| syringe | needle | recovered | dead volume | doses | % 5 doses | % 6 doses | % 7 doses |
| --- | --- | --- | --- | --- | --- | --- | --- |
| 1 ml | 27Gx1/2 | 1,87±0.02 | 0,38±0.02 | 6 | 100 | 100 | 0 |
| 1 ml | 23Gx1 | 1,84±0.02 | 0,41±0.02 | 6 | 100 | 100 | 0 |
| 3 ml | 23Gx1 | 1,69±0.04 | 0,56±0.04 | 5 | 100 | 0 | 0 |
| 3 ml | 27Gx1/2 | 1,83±0.08 | 0,42±0.08 | 5,5 | 100 | 50 | 0 |
| 3 ml | 23Gx1(a) | 2,21±0.05 | 0,04±0.05 | 7 | 100 | 100 | 100 |

**Figure 1.**
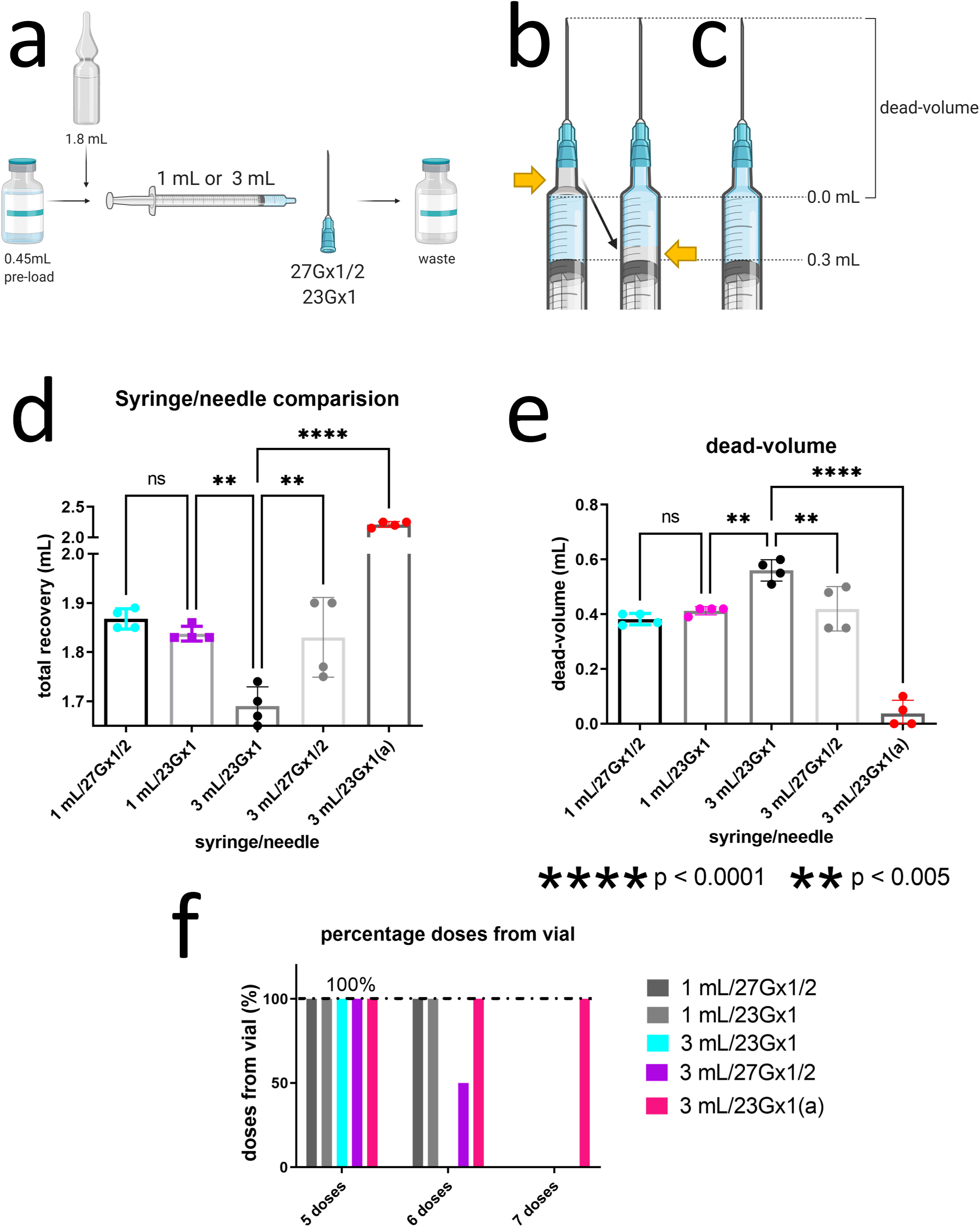
Experimental design and syringe-needle comparison. Schematic of testing process (a); Filled syringe-needle system with and without dead-volume (b). Total recovery of 0.9% Sodium Chloride from a 2.25 mL filled vial using different syringe-needle systems (c); Total dead-volume of 0.9% Sodium Chloride from a 2.25 mL filled vial using different syringe-needle systems (d); Percentage of doses recovered from a 2.25 mL filled vial using different syringe-needle systems (e).

Next, we showed that is possible to use every full dose, the sixth, or even a seventh dose from a multiple-dose vial. We obtained a sixth dose from each vial 50% of the times using a 3ml-27Gx1/2 needle system for the dose extraction (Table 1, Fig. 1f).

Then, after we had stablished 27Gx1 needle is the best option for the doses withdraw, we compared 3mL to 1mL syringes. We identified that 1mL syringe constantly improved the total volume recovered from the multiple-dose vial compared to 3mL (Figure 1d). Also, 1 mL syringe decreased the dead-volume compared to 3 mL syringe (Figure 1e). Moreover, independently of the needle Gauge, 1 mL syringe constantly recovered six doses from the multiple-dose vial compared to 3mL (Figure 1f). On the other hand, there is no differences in the number of doses draw, total volume, or dead volume collected by using 23G or 27G needles attached to a 1mL syringe (Fig. 1d-f).

Finally, we obtained the seventh dose from a multiple-dose vial using a no dead-volume approach (Figure 1b, f).

The data suggest that switching to 1 mL-23G system or to a no dead-volume approach from any of the others tested would provide between 20 and 40% additional vaccine doses per vial if the current 5-dose vials are used (Table 1)

## DISCUSSION

Now, it is our duty, as physicians and nurses, to ensure the efficacy and efficiency of the worldwide COVID-19 vaccination process. However, if standard syringes-needles and technique are used, there may not be sufficient volume to draw extra doses from a single vial. Indeed, we learned this important lesson from the Influenza pandemic^10,17^.

If we only extracted five doses instead of six, as described on the vial label of the first FDA COVID-19 vaccine approved, we could lose up to 1.4 billion doses. In this way, we could waste not only doses, but priceless time, extending the vaccination campaign, billions of dollars, and most importantly, human lives. But not only the high dead-volume inside the syringes and needles could be a problem. In the vaccination practice, and according to the manufacturer ^18^, any further remaining product inside the vial should not be pooled with other vial residue to obtain extra doses.

There is an increasing COVID-19 vaccine demand that we estimate up to 14 billion dual doses. We are on time to advise the public health authorities to make the best possible decision regarding to the syringe systems to face the COVID-19 vaccination process. Remains pending for future studies to test different COVID-19 vaccines with low dead-volume syringe to evaluate the number of doses obtained as previously described ^17^.

## METHODS

This study did not include human subject research, then, institutional review board authorization was not necessary ^16^. We used 0.9% Sodium Chloride, two types of needles and two types of high dead-volume syringes for effectiveness analysis. All experiments in quadruplicate.

### Dose Preparation and dilution

For the procedure involved 1 mL and 3 mL syringes (Nipro syringe) we used a 27 Gauge ½ “needle (0.4 x 13mm) and a 23 Gauge 1” needle (0.6mm x 25mm), all systems used with luer lock.

Four different syringe-needle system were tested (Figure 1a): 1 mL syringe attached to 23- or 27-Gauge needle and 3 mL syringe attached to 23- or 27-Gauge needle (23G or 27G respectively). To quantify the number of microliters recovered from the vials, we calculated the number of syringes filled in with 0.3mL of solution times 0.3 plus the last dose extracted using a 1 mL syringe.

Clean and dry 5 mL-vials capacity (FluQuadri, Sanofi Medley) were used for all experiments. 0.45 mL of 0.9% Sodium Chloride (Baxter, for IV Infusions in Viaflex Container) was loaded in one vial using a 1 mL syringe −23G needle system representing the *undiluted vial*. 1.8 mL 0.9% Sodium Chloride was diluted in the 0.45 mL “undiluted vial” using a 3 mL syringe-23G needle system. We made sure to remove all air prior to drawing up. We used new needles and syringes in each draw, mimicking a vaccination procedure.

### No dead-volume approach

We considered syringe-needle dead space was the volume of residual fluid that remains within the syringe-needle system after the plunger is fully depressed during inoculation^11,16^. A different technique for the no dead-volume approach was used by considering 0.0 mL as the starting point and filled in until 0.3 milliliters (Figure 1b left). After 0.3 mL reached, an extra 0.1-0.2 mL air volume was added to force all fluid draw out the syringe-needle system. The air volume was positioned in the plunger side of the syringe (Figure 1b right).

### Statistical Analysis

The data are presented as the mean ± S.D. We used one-way ANOVA to compare more than two groups. P < 0.05 was considered statistically significant. Tukey’s test for multiple comparison. We used GraphPad Prism 9 for Windows to perform statistical analysis. Graphical representation were created with BioRender.com.

## Data Availability

All data is in table 1

## ACKNOWLEDGEMENTS

The authors are grateful to Gladys Paredes Ulloa, Carlos Castro, Patricia Nilo, Jocelyn Araya, CESFAM Sarmiento and Ilustre Municipalidad de Curico. for technical assistance and financial support. This study was supported, in part, by the Coordenação de Aperfeiçoamento de Pessoal de Nível Superior – Brasil (CAPES) – Finance Code 88882.434714/2019–01.

## CONFLICTS OF INTEREST

No conflicts of interest

## AUTHOR CONTRIBUTIONS

CPJ: Conceptualization, Formal analysis, Methodology, Investigation, Writing - Original Draft, Writing - Review & Editing

LAV: Formal analysis, Writing - Review & Editing, Supervision

EPA: Conceptualization, Methodology, Formal analysis, Resources, Writing - Original Draft, Writing - Review & Editing, Supervision

## Notes

Financial support: Coordination of Improvement of Higher-Level Personnel of Brazil (CAPES)

### Competing Interest Statement

The authors have declared no competing interest.

### Funding Statement

This study was supported, in part, by the Coordenacao de Aperfeicoamento de Pessoal de Nivel Superior Brasil (CAPES) Finance Code 88882.434714 2019 01.

### Author Declarations

This study did not include human subject research, then, institutional review board authorization was not necessary

